# Developing intrinsic capacity measures for research and practice in an Australian context

**DOI:** 10.64898/2026.07.23.26358833

**Authors:** Jiayue Wang, Philip Clare, Vasi Naganathan, Ding Ding, Matteo Cesari, Stéphane Cullati, Julie Byles, Fiona M. Blyth, Saman Khalatbari-Soltani

**Author notes:** **Corresponding author:** Dr Saman Khalatbari-Soltani, School of Public Health, Faculty of Medicine and Health, The University of Sydney, New South Wales, Australia. Co-first author (Jiayue Wang, Philip Clare).

## Abstract

**Introduction:** Intrinsic capacity is a key construct for healthy ageing; however, its measurement remains heterogeneous, with insufficient attention to context, data availability, and intended purpose. We operationalised intrinsic capacity to facilitate its assessment in both research and clinical settings.

**Method:** Using data from Concord Health and Ageing in Men Project (CHAMP; 2005-2007; 1,705 Australian men aged ≥70 years), we developed three intrinsic capacity models using confirmatory factor analysis (CFA): literature-based (21 commonly used indicators), minimum-set (10 indicators), and clinically informed (10 indicators selected for clinical feasibility). Convergent validity was assessed using activities of daily living (ADL) and instrumental activities of daily living (IADL) as indicators of functional ability. Predictive validity was examined using 2-year incident ADL and IADL dependency and 10-year mortality follow-up data.

**Results:** All models demonstrated acceptable model fit (robust root mean square error of approximation <0.06). The second-order level of CFA showed significant associations with the cognition, locomotor function, psychological well-being, and vitality domains, but not the sensory domain. Higher intrinsic capacity scores were associated with lower ADL (e.g., IC_clinically informed_ 0.91 [0.90-0.92]) and IADL dependence (e.g., IC_clinically informed_ 0.92 [0.92-0.93]) at baseline, lower incident dependency at follow-up, and lower all-cause mortality (e.g., IC_minimum-set_ 0.96 [0.96-0.97]), demonstrating convergent and predictive validity.

**Conclusion:** This study developed and evaluated three intrinsic capacity models to address measurement challenges and intended use, supporting a validated yet flexible approach to measuring intrinsic capacity in research and practice. Their adoption across diverse research and clinical contexts would facilitate comparability and strengthen the evidence base.

**Key Points:**

- Intrinsic capacity, the composite of an individual’s physical and cognitive capacities, is a key construct for healthy ageing. Purpose-built operationalisations of intrinsic capacity tailored to research and clinical contexts are needed.
- Three intrinsic capacity models were developed addressing current challenges related to data availability, measurement feasibility, and intended use, all demonstrating good model fit and convergent and predictive validity.
- A clinically informed model developed with geriatrician input demonstrated that a parsimonious set of indicators can perform comparably to a comprehensive literature-based model, without compromising validity.
- Adoption of these models across diverse populations and settings would strengthen the evidence base and facilitate comparability, ultimately informing policy and practice.

## Introduction

The global population is ageing rapidly [1]. The proportion of the world’s population aged 65 and over is predicted to reach 38% in 2050, more than three times the 11% recorded in 1950 [2]. However, if the extra years of life are dominated by a rapid decline in physical and mental capacities, older people’s ability to participate fully in life will be limited, with negative implications for them and broader society. In the context of this demographic shift, maximising the period individuals spend in a state of health [3] and minimising the period of living in poor health or with disability (i.e. compression of morbidity) are pressing contemporary public health challenges.

Healthy ageing, as defined by the World Health Organization (WHO) in 2015, refers to the process of developing and maintaining an individual’s functional ability that enables wellbeing in older age, marking a shift away from a focus on the presence or absence of disease [4]. Functional ability encompasses the capabilities that enable people to be and to do what they have reason to value. It consists of intrinsic capacity, environmental characteristics, and the interaction between them [4]. Intrinsic capacity reflects the composite of an individual’s physical and cognitive capacities and is generally considered to include five domains: cognition, vitality, locomotor function, sensory capacity, and psychological well-being [5–7].

Despite a growing recognition of the need to assess older people’s health in a multidimensional way and the effort to measure healthy ageing across diverse populations and contexts, a 2023 review highlighted that the measurement of intrinsic capacity remains inconsistent, reflecting a lack of a standardised approach to define and operationalise it [8, 9]. The field is still progressing toward consensus on the conceptual definition of each domain; evidence on which specific indicators best represent them remains limited and evolving; and the heterogeneity of measures used across studies is further driven by gaps in availability of relevant data across existing datasets [5, 7]. This heterogeneity poses challenges for interpreting and comparing evidence across studies and hinders progress towards developing and adopting standardised approaches to intrinsic capacity assessment in clinical practice, public health monitoring, and policy planning [10]. Achieving a fully standardised approach to intrinsic capacity measurement therefore remains a work in progress, contingent on advances in both scientific consensus and data availability [5, 9].

Intrinsic capacity measures serve different purposes: comprehensive measures are needed in research to build the evidence base on domain indicators, while brief measures, replicable across diverse settings and datasets, are needed for population health monitoring and clinical practice. Given the current gaps in data availability and the need for comparability of evidence across studies, there is a case for developing parsimonious measures that best represent intrinsic capacity using a limited number of indicators. In clinical practice, brief, evidence-derived measures that are feasible for routine use are particularly needed to identify older adults at risk of functional decline without placing burden on clinicians or patients, consistent with the complementary roles of intrinsic capacity and frailty along a continuum of care from prevention and health promotion to complex care management [11].

One strategy to address these measurement challenges is to develop and compare multiple operationalisations of intrinsic capacity that vary in their complexity and data requirements, reflecting the different needs of researchers, public health practitioners, and clinicians. In this study, we developed and compared three operationalisations of intrinsic capacity, for the first time in an Australian context, using data from a population-based cohort study of older Australian men. Specifically, we examined: a literature-informed model based on commonly-used domain-specific indicators to facilitate comparability with previous studies; a minimum-set model based on the best performing indicators from the literature-informed model, that explicitly accounted for common challenges in operationalising a multidimensional construct, including limited data availability; and a clinically-informed model that evaluated the feasibility, for routine clinical settings, of measures commonly used in cohort studies.

## Method

### Study description

We used data from the Concord Health and Ageing in Men Project (CHAMP), a longitudinal observational study of 1705 ethnically diverse men aged 70 years and over living in three local government areas near Concord Hospital in Sydney, New South Wales, Australia (mean age at baseline 77 years) [12]. Baseline data collected between 2005 and 2007 were used to develop models to operationalise intrinsic capacity. First follow-up data collected between 2007 and 2009 and linked mortality data up to December 2017 were used to validate the three developed intrinsic capacity constructs (Figure S1) [13]. Our study methods were reported in accordance with the Strengthening the Reporting of Observational Studies in Epidemiology (STROBE) guidelines (Table S1) [14].

### Measures

#### Intrinsic capacity domains

The inclusion of commonly used domains and related indicators was informed by a review of the available literature, mainly from related systematic reviews and WHO reports/guidelines [5, 9, 15–17]. The five domains of intrinsic capacity, cognition, locomotor function, vitality, sensory capacity, and psychological well-being, were consistent with those proposed by the WHO [5, 18].

#### Indicators

The inclusion of indicators for each domain was guided by available literature and data availability, with details of their measurement methods and variable categorisation presented in Supplementary Materials Appendix 1 and Table S2. Briefly, we considered: Mini-Mental State Examination (MMSE) [19], verbal fluency [20], Trail Making Test (TMT) [21], Weigl Color Form Sorting Test [22], immediate logical memory, and delayed logical memory by the Wechsler Memory Scale [23] for cognition; forced expiratory volume in 1 second (FEV1), hand grip strength, body mass index (BMI), weight loss [24], and waist-to-hip ratio (WHR) [25] for vitality; walking pace, postural sway, 20-centimetre narrow walk, single and repeated chair stand, and knee strength for locomotor function; hearing loss and visual acuity for sensory capacity; and depression [26], social interaction, social satisfaction [27], anxiety [28], self-reported health worries, and self-reported poor sleep for psychological well-being [28]. Given inconsistency in the literature and expert advice, we considered hand grip strength as an indicator of both vitality and locomotor domains.

#### Other measurements

To describe baseline characteristics of the participants, we considered demographic information, including age (continuous), country of birth (Australian born, yes/no), marital status (single, married/partnered, widowed/separated/divorced), living arrangement (living alone, yes/no), and socioeconomic position. Socioeconomic position was assessed using four separate indicators: highest education level, occupation category, source of income, and home ownership (owning home outright versus other arrangements, such as leasing or purchasing in a retirement village, paying rent to a private landlord, and paying rent to the government for public housing) (see Appendix 2 for details).

To assess the convergent and predictive validity of the three developed intrinsic capacity constructs (details provided in the section below), the outcome measures included Activities of Daily Living (ADL) [29] and Instrumental Activities of Daily Living (IADL) as indicators of functional ability [5, 30]. ADL captures basic self-care tasks (e.g., bathing), whereas IADL captures more complex tasks required for independent living (e.g., managing finances). ADL was assessed using seven questions adapted from the Katz ADL scale for use in the general population, asking whether the participant required help from another person or the use of special equipment to perform each activity [29]. Participants who face any difficulty or require any help with the ADLs are defined as being ADL dependent [31]. IADLs were assessed by the Older Americans Resources and Services (OARS) Instrumental Activities of Daily Living [30], which contains eight tasks. IADL dependency was defined as the need for any assistance with one or more IADL tasks [32]. Detailed information on ADL and IADL can be found in Table S3.

We also used all-cause mortality to assess the predictive validity of the three developed intrinsic capacity constructs, with the mortality data acquired from the data linkage to the NSW Registry of Births, Deaths, and Marriages and the Australian Bureau of Statistics/Australian Coordinating Registry Cause of Death Unit Record File up to December 2017 [13].

### Statistical analysis

#### Handling missing data

We included all baseline participants in CHAMP (n = 1705) (Figure S1). Among these, 606 participants had complete data for all intrinsic capacity indicators, and an additional 723 participants had up to two missing indicators. The proportion of missing data was below 20% across most indicators used in developing the intrinsic capacity measure, except for FEV1 (24.2%) and knee strength (26.2%) (Table S2).

Missing data due to non-response and non-responders were addressed using a multiple imputation by chained equations (MICE) model with random forests [33], with 27 imputations performed (see Appendix 3 for further information).

#### Development of the intrinsic capacity construct

We use confirmatory factor analysis (CFA) to develop the intrinsic capacity construct under the structural equation modelling framework [5, 34]. To ensure consistency in interpretation, some continuous indicators were reverse-scaled so that higher values uniformly reflected better capacity or performance, and some binary indicators were recoded such that a value of 1 indicates better capacity (see Appendix 4 for details). The initial literature-based CFA model included 21 commonly used indicators across the intrinsic capacity domains and available from CHAMP data. The model was specified as a hierarchical structure, with first-order latent factors capturing the observed within-domain indicators and second-order latent factors representing each domain of intrinsic capacity (cognition, vitality, locomotor function, sensory capacity and psychological well-being), following the healthy ageing framework proposed by the WHO [5, 18]. The covariance structure was pre-specified by allowing residuals of indicators measured using the same assessment tool to correlate, accounting for method specific variance. The variances of the latent variables were standardised to 1 to enhance the interpretability of the model and the predicted intrinsic capacity score.

Given that one aim of the study was to develop a construct of intrinsic capacity that could be accessible and easily implemented in healthcare settings, and to address the challenge of limited data availability across diverse datasets, we subsequently tested a series of simplified models to develop a minimum-set model. These models were developed by reducing the number of indicators included in each domain, using several different approaches. First, we excluded indicators with the lowest factor loading within each domain since they define the construct more weakly than indicators with higher loadings [35, 36], while ensuring that each domain maintained at least two indicators (the minimum required for the latent factor structure to remain identifiable and to preserve the multidimensional nature of each domain) [37]. Second, we excluded indicators with the highest standard error within each domain, because a high standard error indicates low precision [35]. Third, we tested models that included, where applicable, the three indicators with the highest factor loading from each domain of the full model. Fourth, we evaluated each domain separately by fitting separate CFA models for each of the five domains, in which indicators were selected based on having the highest two or three factor loadings, the lowest two or three standard errors, or a combination of both. The selected indicators from each of the five domain-specific models were then combined into a single CFA model representing all five domains. Throughout the indicator selection process, we also considered the distinctiveness and content coverage of the indicators, ensuring that each domain was represented by measures capturing various aspects of the construct. In addition, to develop a clinically informed model, guided by the geriatrician’s recommendations, we tested additional models prioritising indicators with greater clinical utility, including commonly used indicators and those without requiring specialised equipment from the initial full model (Appendix 5).

For all models, model fit was assessed using a set of goodness-of-fit statistics, including robust root mean square error of approximation (RMSEA), comparative fit index (CFI), Tucker–Lewis index (TLI), and standardised root mean square residual (SRMR). A RMSEA value below 0.06, CFI and TLI above 0.95, and SRMR below 0.08 are generally considered to indicate a relatively good model fit [38]. Among these, robust RMSEA was prioritised as the primary criterion for model selection because it accounts for model complexity and is less sensitive to sample size than other fit indices, with CFI and TLI additionally considered and reported to complement the evaluation of model fit.

The three models were used to estimate participant-level intrinsic capacity. To enhance interpretability, scores were rescaled to a range of 0 to 100, where a higher score represents better intrinsic capacity; details of rescaling and the scoring formula are available in Appendix 4.

We also conducted item response theory (IRT) analysis to evaluate the discriminatory ability of the indicators for measuring each domain of intrinsic capacity. Further details on the IRT modelling are included in Appendix 6.

#### Convergent validity

We used the intrinsic capacity scores estimated from the three CFA models to assess convergent validity cross-sectionally by examining their association with baseline functional ability, measured using ADL and IADL [39], which serve as independent measures of functional ability, assessed separately from the indicators included in constructing intrinsic capacity [15]. They are commonly used in the literature, allowing comparison with existing studies, and have demonstrated the highest discrimination for capturing information on intrinsic capacity and functional ability [7].

#### Predictive validity

We assessed the predictive validity of the intrinsic capacity scores at baseline in identifying the onset of functional dependence at first follow-up (mean follow-up years of 2.16; SD = 0.2), among participants who were ADL or IADL independent at baseline. Functional dependence was defined as requiring help with at least one ADL or IADL item, and functional independence as reporting no such need for help at baseline. Baseline intrinsic capacity scores were regressed on functional dependence status at first follow-up using logistic regression, with estimates presented as odds ratios (ORs) and 95% confidence intervals (CIs).

Additionally, a Cox proportional hazard model was fitted to further evaluate the predictive validity of the intrinsic capacity structure, with time to all-cause mortality as the outcome and the baseline intrinsic capacity score as the predictor, to examine the association between intrinsic capacity and mortality risk. Hazard ratios (HRs) and 95% confidence intervals (CIs) were reported. Survival time was measured as the time from the date of baseline interview to either the date of death, or the end of follow-up (December 31, 2017 for all-cause mortality).

## Results

At baseline, most participants were married/partnered (76.8%), about half were born in Australia (49.8%), and the majority owned their home outright (87.6%). The sociodemographic characteristics and baseline scores for each of the included indicators across all domains are shown in Table 1.

**Table 1.** Characteristics of participants at baseline, the CHAMP study (n = 1705)

| **Characteristics** | **Total study population**<br>**(n=1705)** |
| --- | --- |
| **Sociodemographic characteristics** |  |
| Age, years, Mean (SD) | 77 (5.5) |
| Marital status, n (%) |  |
| Single | 86 (5.0) |
| Married or partnered | 1309 (76.8) |
| Widowed separated, or divorced | 310 (18.2) |
| Living alone, yes, n (%) | 318 (18.7) |
| Australian born, n (%) | 849 (49.8) |
| Highest education <sup>a</sup>, n (%) |  |
| High | 206 (12.1) |
| Intermediate | 705 (41.3) |
| Low | 777 (45.6) |
| House ownership status, yes, n (%) | 1494 (87.6) |
| Occupational level <sup>b</sup>, n (%) |  |
| High | 505 (29.6) |
| Intermediate | 627 (36.8) |
| Low | 562 (32.9) |
| Sources of income, n (%) |  |
| Age pension only | 668 (39.2) |
| Age pension and other | 270 (15.8) |
| Other only | 742 (43.5) |
| **Intrinsic capacity domains** |  |
| **Cognition** |  |
| MMSE score, Mean (SD) | 27 (3.05) |
| Logical memory score, Mean (SD) | 10 (4.39) |
| Logical memory recall score, Mean (SD) | 8 (4.35) |
| Colour form sort, n (%) |  |
| Unable to do the test | 88 (5.2) |
| Unable to do the first sort | 464 (27.2) |
| Sorts one category spontaneously | 1007 (59.1) |
| Sorts two categories spontaneously | 3 (0.2) |
| TMT-B, completed in 5 minutes, n (%) | 1007 (59.1) |
| Verbal fluency, Mean (SD) | 9 (2.88) |
| **Vitality** |  |
| Weight loss <sup>c</sup>, yes, n (%) | 129 (7.6) |
| FEV1, Mean (SD) | 2 (0.6) |
| Handgrip strength, Mean (SD) | 35 (7.67) |
| BMI <sup>d</sup>, Mean (SD) | 28 (4.03) |
| Waist-to-hip ratio, Mean (SD) | 1 (0.05) |
| **Locomotor function** |  |
| 6-meter walking pace, (m/s), Mean (SD) | 1 (0.2) |
| 20-cm narrow walk, n (%) |  |
| Did test & had $\leq 2$ deviations at least once | 1197 (70.2) |
| Did at least one trial but $\geq 3$ deviations on all attempts | 415 (24.3) |
| Unable to do any trials | 70 (4.1) |
| Repeated chair stand, time in seconds, Mean (SD) | 16.6 (5.9) |
| Single chair stand, n (%) |  |
| Stands without using arms | 1561 (91.6) |
| Rises using arms | 44 (2.6) |
| Unable to stand | 36 (2.1) |
| Floor sway area, Mean (SD) | 5.08 (6.8) |
| Foam sway area, Mean (SD) | 11.8 (19.3) |
| Leg strength, Mean (SD) | 29 (7.7) |
| **Sensory capacity** |  |
| Hearing loss, yes, n (%) | 1027 (60.2) |
| Visual acuity, Mean (SD) | 1 (0.3) |
| **Psychological wellbeing** |  |
| Depressive symptoms, yes, n (%) | 246 (14.6) |
| Social interaction, Mean (SD) | 8.81 (1.5) |
| Social satisfaction, Mean (SD) | 19 (2.4) |
| Social network <sup>e</sup> |  |
| None | 58 (3.4) |
| 1-2 people | 199 (11.7) |
| More than 2 people | 1415 (83.0) |
| Anxiety, yes, n (%) | 123 (7.2) |
| Sleep problems, yes, n (%) | 470 (27.6) |
| Health worries, yes, n (%) | 424 (24.9) |
| **Measure of functional ability** |  |
| ADL dependent, yes, n (%) | 141 (8.3) |
| IADL dependent, yes, n (%) | 697 (40.9) |

Sample sizes vary due to missing data.
Abbreviations: SD, standard deviation; MMSE, Mini-Mental State Examination; TMT, Trail Making Test; FEV1, forced expiratory volume in 1 second; BMI, body mass index; ADL, activities of daily living; IADL, instrumental activities of daily living.
<sup>a</sup> Education level categories: High, university degree; Intermediate, trade, apprenticeship, certificate, or diploma; Low, no post-school qualification.
<sup>b</sup> Occupation level categories: High, managers/professionals; Intermediate, technicians, trade workers, personal service, clerical & administrative; Low, sales, machinery, labourers.
<sup>c</sup> Weight loss is defined as 15% lower than self-reported heaviest weight (or weight at 25 if the heaviest is missing).
<sup>d</sup> BMI was calculated using height and weight measured during clinical assessment.
<sup>e</sup> Number of persons within one hour travel from the participant's home that the participant feels can depend on or feels very close to.
Details on measurements are provided in Appendices 1 and 2.

### Intrinsic capacity constructs: CFA and IRT

The initial full CFA model (i.e. literature-based model) with 21 indicators demonstrated moderate fit (robust RMSEA=0.026) (Table S4). The path diagram including factor loadings of each indicator is shown in Figure 1. Within the cognition domain, all indicators showed strong factor loadings. In the vitality domain, FEV1 showed strong but not statistically significant factor loadings, while handgrip strength and BMI showed less meaningful associations with the latent vitality domain. Within the locomotor function domain, walking pace, leg strength, repeated chair stand, and narrow walk test demonstrated relatively strong factor loadings, while sway measures and single chair stand showed modest loadings.

**Figure 1.**
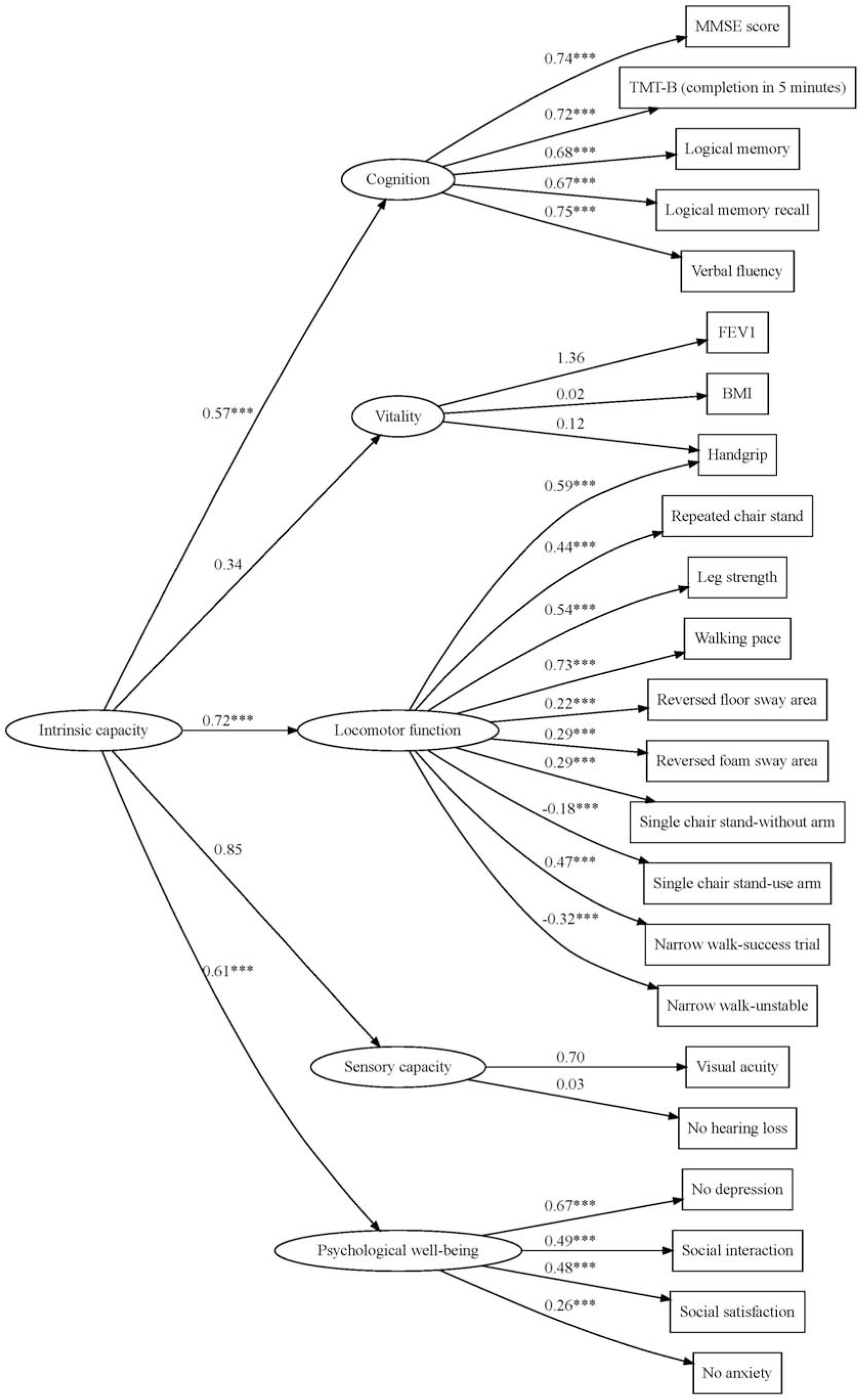
Path diagram of the full confirmatory factor analysis model (Literature-based model) of developing intrinsic capacity construct Abbreviations: MMSE, Mini-Mental State Examination; TMT, Trail Making Test; FEV1, forced expiratory volume in 1 second; BMI, body mass index. Note: details of each indicator are available in Supplementary Materials Appendix 1 and Table S1.sterisks (***) on the paths indicate statistically significant factor loadings (p < 0.001). Numbers shown on the paths represent standardised factor loadings.

Allowing handgrip strength to cross-load on both vitality and locomotor domains resulted in a strong loading on the locomotor domain, with little evidence of association with the vitality domain. Both indicators in the sensory capacity domain, visual acuity and hearing loss, showed low, non-significant factor loadings. Within the psychological well-being domain, social interaction, social satisfaction, absence of depression and absence of anxiety all showed significant factor loadings. At the second-order level of the CFA model, intrinsic capacity was significantly associated with cognition, locomotor function and psychological well-being. The vitality and sensory capacity domains showed a moderate second-order factor loading.

Based on extensive comparisons across a series of model examinations and evaluation of model fit indices, a minimum-set model retaining two indicators per domain was identified (Figure 2 and Table S4). MMSE and TMT-B represented cognition; FEV1 and hand grip strength represented vitality, following the removal of BMI due to its consistently low factor loading; repeated chair stand and walking pace represented locomotor function; and social interaction and the absence of depression represented psychological wellbeing. Despite hearing showing a weaker factor loading than vision, both were retained in the sensory domain as they represent distinct sensory capacities and were the only available sensory measures in CHAMP. This simplified minimum-set model demonstrated good fit (robust RMSEA=0.024).

**Figure 2.**
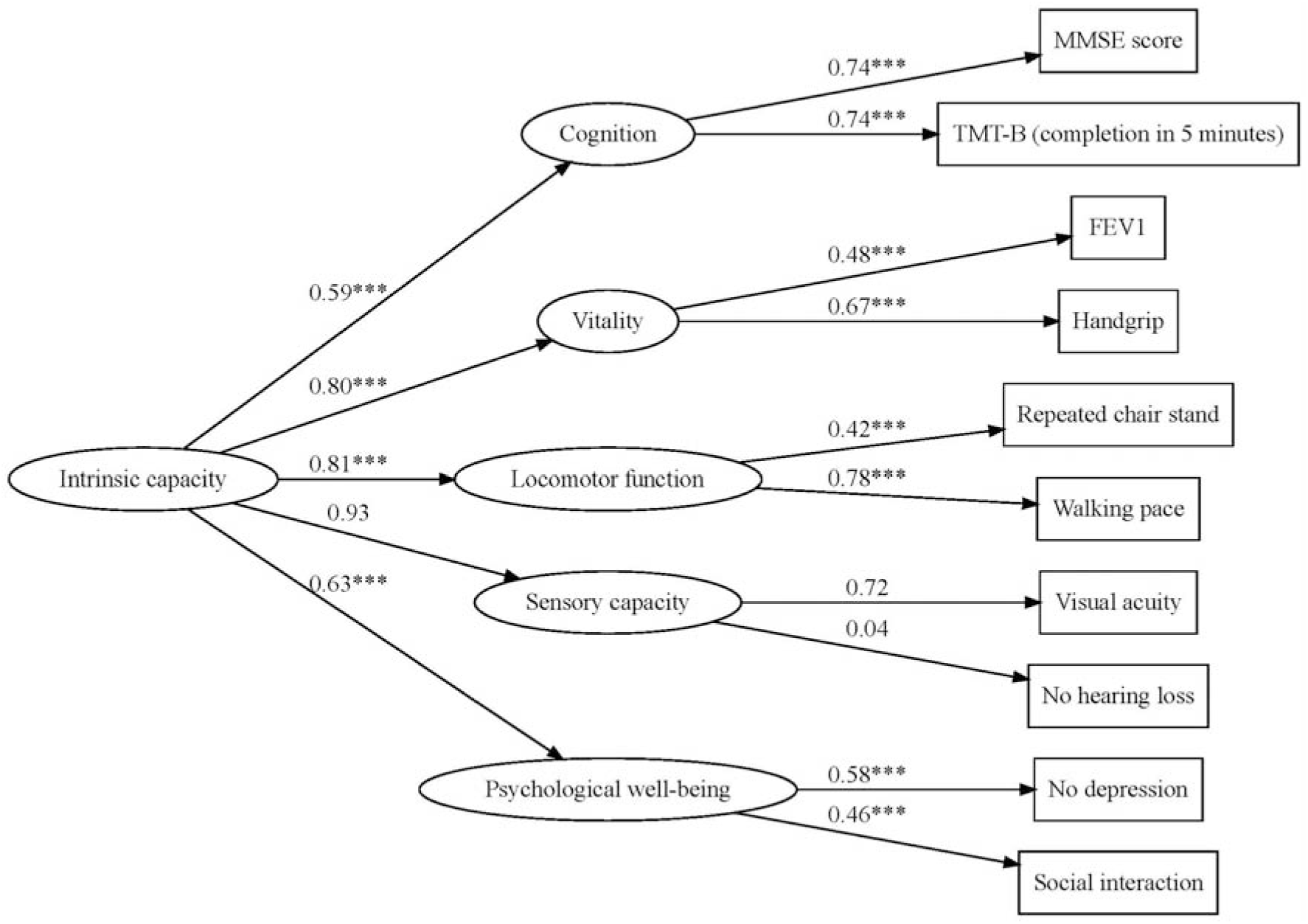
Path diagram of the simplified confirmatory factor analysis model (Minimum-set model) of developing intrinsic capacity construct. Abbreviations: MMSE, Mini-Mental State Examination; TMT, Trail Making Test; FEV1, forced expiratory volume in 1 second. This simplified model was selected based on superior model fit and construct validity. Asterisks (***) on the paths indicate statistically significant factor loadings (p < 0.001). Numbers shown on the paths represent standardised factor loadings.

To develop a clinically informed model, alternative models were further evaluated in which indicator selection was guided by recommendations from a geriatrician, prioritising more feasible measures in health care settings. The clinically informed model included: logical memory and MMSE for cognition; handgrip strength and FEV1 for vitality; repeated chair stand and walking pace for locomotor function; hearing loss and visual acuity for sensory capacity; and absence of depression and absence of anxiety for psychological well-being (Figure 3). This model demonstrated good overall fit (robust RMSEA=0.019). Given that FEV1 require specialised equipment and may not be available in all clinical settings (Table S4, models 23-24), we additionally evaluated a model excluding this indicator. This model retained an acceptable fit (robust RMSEA=0.020).

**Figure 3.**
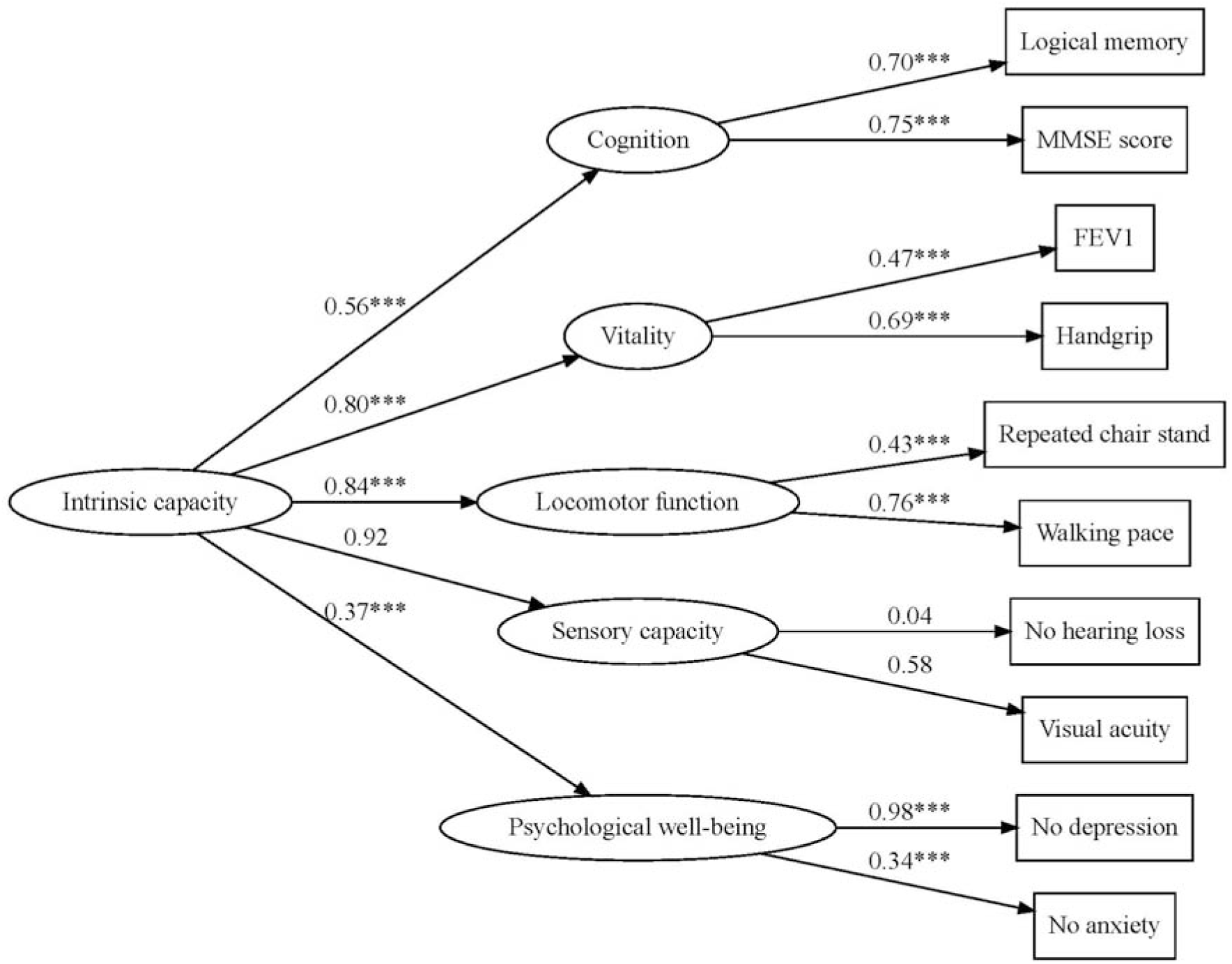
Path diagram of the clinically informed confirmatory factor analysis model (Clinically informed model) of developing intrinsic capacity construct Abbreviations: MMSE, Mini-Mental State Examination; FEV1, forced expiratory volume in 1 second. The clinically informed model assessed the feasibility, for routine clinical settings, of complex and time-consuming measures commonly used in cohort studies. Asterisks (***) on the paths indicate statistically significant factor loadings (p < 0.001). Numbers shown on the paths represent standardised factor loadings.

The IRT analyses supported the findings from the CFA models. Indicators that demonstrated stronger factor loadings in the CFA generally showed higher discrimination in the IRT models (Appendix 6, Figure). These findings provide additional evidence supporting the selection of indicators in the minimum-set model and the need for more than one indicator to better capture the full spectrum of each intrinsic capacity domain.

### Convergent validity and predictive validity of three intrinsic capacity constructs

Mean (SD) intrinsic capacity scores were 61.74 (15.25) for the literature-based model, 55.65 (15.58) for the minimum-set model, and 54.47 (15.24) for the clinically informed model (Figure S2). Higher intrinsic capacity scores were significantly associated with lower odds of ADL and IADL dependence cross-sectionally at baseline using all three models (Table 2), demonstrating convergent validity. In prospective analyses, higher baseline intrinsic capacity scores were significantly associated with lower odds of functional dependence at first follow-up and a lower risk of all-cause mortality over a mean (SD) follow-up of 9.11 (SD 3.54) years (Table 3), demonstrating predictive validity across three models.

**Table 2.** Convergent validity – cross-sectional associations between the estimated intrinsic capacity scores and measures of functional ability.

|  | ADL |  |  | IADL |  |  |
| --- | --- | --- | --- | --- | --- | --- |
|  | Independent | Dependent |  | Independent | Dependent |  |
| Intrinsic capacity score | mean (SD) | mean (SD) | OR (95% CI) | mean (SD) | mean (SD) | OR (95% CI) |
| **Literature-based model<sup>a</sup>** | 63.47 (14.17) | 42.53 (13.60) | 0.91 (0.90-0.92) | 67.76 (11.82) | 53.32 (15.52) | 0.93 (0.92-0.93) |
| **Minimum-set model<sup>b</sup>** | 57.27 (14.63) | 35.56 (12.89) | 0.91 (0.90-0.92) | 61.72 (12.57) | 47.17 (15.43) | 0.93 (0.92-0.94) |
| **Clinically informed model<sup>c</sup>** | 56.15 (14.32) | 35.80 (12.31) | 0.91 (0.90-0.92) | 60.51(2.29) | 46.02 (14.95) | 0.92 (0.92-0.93) |

Abbreviations: ADL, activities of daily living; IADL, instrumental activities of daily living; OR, odds ratio; CI, confidence interval.
Table reports means and standard deviations of intrinsic capacity by ADL group, and tests odds of being ADL or IADL dependent associated with a 1-unit increase in intrinsic capacity.
ORs were obtained from the univariable logistic model of ADL/IADL dependence on the intrinsic capacity score.
Note: The estimated intrinsic capacity scores were normalised and rescaled to a range from 0 to 100.
<sup>a</sup> Literature-based model: the confirmatory factor analysis (CFA) model including all 21 baseline indicators across the five domains of intrinsic capacity.
<sup>b</sup> Minimum-set model: the CFA model retaining the reduced set of 10 baseline indicators.
<sup>c</sup> Clinically informed model: the CFA model containing 10 baseline indicators, developed to enhance feasibility and applicability in healthcare settings.

**Table 3.** Predictive validity – longitudinal associations between baseline estimated intrinsic capacity scores and subsequent incident ADL/IADL and all-cause mortality.

|  | **Incidence of ADL dependence** | **Incidence of IADL dependence** | **All-cause mortality** |
| --- | --- | --- | --- |
| **Intrinsic capacity score** | **OR (95% CI)<sup>d</sup>** | **OR (95% CI)<sup>d</sup>** | **HR (95% CI)<sup>e</sup>** |
| **Literature-based model<sup>a</sup>** | 0.92 (0.91-0.94) | 0.94 (0.93-0.95) | 0.96 (0.96-0.97) |
| **Minimum-set model<sup>b</sup>** | 0.92 (0.91-0.94) | 0.94 (0.93-0.96) | 0.96 (0.96-0.97) |
| **Clinically informed model<sup>c</sup>** | 0.92 (0.90-0.93) | 0.94 (0.93-0.96) | 0.96 (0.96-0.97) |

Abbreviations: ADL, activities of daily living; IADL, instrumental activities of daily living; OR, odds ratio; CI, confidence interval.
<sup>a</sup> Literature-based model: the CFA model including all 21 baseline indicators across the five domains of intrinsic capacity
<sup>b</sup> Minimum-set model: the CFA model retaining the reduced set of 10 baseline indicators
<sup>c</sup> Clinically informed model: the CFA model containing 10 baseline indicators, developed to enhance feasibility and applicability in healthcare settings.
<sup>d</sup> Odds ratios (95% CI) obtained from the univariable logistic model of Wave 2 incident ADL/IADL dependence on the intrinsic capacity score, restricted to participants who were ADL/IADL independent at baseline.
<sup>e</sup> Hazard ratios (95% CI) from the univariable Cox Proportional Hazards models of all-cause mortality.

## Discussion

This study is the first to develop and evaluate intrinsic capacity models using multiple strategies: a literature-based model to facilitate comparability with previous studies; a minimum-set model to address common challenges in operationalising a multidimensional construct, including limited data availability; and a clinically informed model prioritising measures suitable for routine clinical settings over more complex and time-consuming measures commonly used in research cohort studies. Across all three models, intrinsic capacity demonstrated convergent and predictive validity, with higher scores associated with a lower odds of ADL and IADL dependence, and lower risk of all-cause mortality.

Since 2015, efforts have been made to develop multidimensional measures of intrinsic capacity based on the WHO healthy ageing framework across different settings, but none have been conducted in Australia [5, 7, 9, 15, 16, 39–42]. Despite considerable heterogeneity in the domains and indicators included, most developed measures have demonstrated convergent validity and predictive validity for outcomes such as ADL, IADL, and all-cause mortality, consistent with the findings of the present study [15, 39–42]. This study advances the field through a purpose-driven approach, directly responding to the absence of an agreed operationalisation method for intrinsic capacity, a gap that reflects variation in data availability across studies and datasets [43], the age-dependent nature of indicators used in ageing cohorts, and differences in intended use across research and clinical contexts. By comparatively examining alternative indicator combinations and reporting model fit across all iterations, this study provides empirical evidence to inform decisions about how intrinsic capacity can be operationalised under different data and practical constraints.

Our findings demonstrate that a parsimonious set of well-chosen indicators can perform comparably to a larger set, and that different modelling approaches yield measures suited to different purposes. Reporting model fit across all iterations provides empirical support for considering alternative indicators where data availability or clinical constraints exist.

However, further evidence is needed to validate these alternatives across diverse populations and settings, particularly among women and lower-income countries. These findings highlight the need for context-sensitive approaches to developing intrinsic capacity measures that can be adapted to diverse data sources and settings.

All three models supported the multidimensional structure of intrinsic capacity proposed by the WHO, adding to the growing evidence of its applicability across diverse national contexts. At the domain level, cognition, vitality, locomotor function, and psychological wellbeing performed largely as expected based on existing literature, with indicators demonstrating theoretically consistent associations with their respective latent constructs. Within the cognition domain, memory and executive function measures (logical memory, TMT-B, verbal fluency, and MMSE score), showed strong associations, consistent with available literature [16, 44–46]. In the vitality domain, grip strength and respiratory function performed as expected, while BMI, which was objectively measured, showed consistently weak associations, a finding that may reflect the limited sensitivity of BMI as an indicator of vitality in this study context [16, 47]. Within the locomotor function domain, most indicators demonstrated significant association with the latent construct, consistent with previous studies [16, 48–50]. However, repeated chair stand, the narrow walk test, and walking pace showed the strongest and most consistent factor loadings across models, suggesting these measures most reliably capture locomotor functioning. Psychological well-being was reflected by measures of anxiety, depression, and social functioning across the models, though variability in factor loadings across models reflects the inherent breadth of this domain, consistent with previous observations that psychological functioning is among the most heterogeneously measured domains of intrinsic capacity [16].

The sensory domain was an exception: hearing showed consistently weak associations across all three models, and neither hearing loss nor visual acuity showed a meaningful relationship with the sensory latent construct, contrasting with previous evidence [5, 9, 16]. First, hearing in CHAMP was assessed using a single yes/no question, limiting its sensitivity. Second, vision and hearing represent distinct sensory modalities that do not necessarily decline together, and a single latent sensory factor may not adequately capture this heterogeneity.

Third, the use of corrective devices such as glasses or hearing aids may partially compensate for impairment, influencing how sensory limitations are reported or observed [51]. Future studies should consider broader sensory assessments to better capture this domain.

### Strengths and limitations

This study used a sample of older Australian men, with a wide range of objectively measured and self-reported indicators relevant to intrinsic capacity. Indicator selection for the full model was informed by the existing literature, supporting comparability with previous studies.

A key strength is the comprehensive and transparent evaluation of multiple models, considering statistical performance, data availability, model fit, clinical feasibility, and geriatrician input. This approach provides alternative model specifications that can be selected according to context, feasibility, and available data.

There are several limitations to this study. CHAMP is limited to men aged 70 years or older, restricting applicability of our findings to women and younger populations, and highlighting the need for studies examining intrinsic capacity from mid-life onwards. As with most ageing cohort studies, CHAMP is susceptible to selection bias, as participants represent a relatively healthier subset of the older population. Sensory domain indicators were limited to visual acuity and hearing in CHAMP, though these are among the most commonly used measures, facilitating comparability but also underscoring the need for broader sensory measurements in future studies. Indicator selection was guided by existing literature and the WHO definition, though further studies should explore a broader range of indicators, including those not yet well represented in the literature, as limited evidence does not imply limited relevance. In addition, as the field has yet to reach consensus on which indicators best represent each domain, some theoretically important indicators may not have been included. Some self-reported indicators may be subject to measurement or recall bias; however, self-report is widely used in population-based ageing research. The culturally and linguistically diverse composition of CHAMP, combined with use of English-language instruments, may have introduced language- and culture-related bias, potentially affecting both objective and subjective assessments. The intrinsic capacity models were derived from cross-sectional data. As measurement protocols in ageing cohort studies are often adapted over time to reflect declining capacities, the feasibility and comparability of the same indicator set may be affected in longitudinal application. Finally, further studies are needed to cross-compare models developed in this study across different databases, testing their external validity.

### Conclusions

We developed and evaluated three intrinsic capacity models to address challenges related to data availability, measurement feasibility, and application across research and clinical settings. By integrating evidence from the literature with statistical evaluation and clinical input, the models provide adaptable approaches to measuring intrinsic capacity. Together, they support a more standardised yet flexible framework that facilitates comparability across studies while accommodating diverse contexts, with the parsimonious approach in particular potentially offering a cost-effective option that can be adapted to local resources and settings. Although developed in a sample of older men, these findings provide an empirical basis for operationalising intrinsic capacity, offering a starting point for further validation across diverse populations and settings.

## Ethical Approval and Consent to Participate

The CHAMP study complied with the World Medical Association Declaration of Helsinki and was approved by the Sydney South West Area Health Service Human Research Ethics Committee. Written informed consent was obtained from all participants involved in the study.

## Data availability

Some access restrictions apply to the data underlying this study’s findings. The original human ethics committee approval for the Concord Health and Ageing in Men Project (CHAMP) in 2004 did not allow for data to be sent outside Australia. Furthermore, the participants in CHAMP have not consented to their data being distributed beyond the CHAMP investigators and their associates. Qualified researchers may submit a request to the CHAMP Management Committee and access will require additional ethics approval from the Sydney LHD HREC - CRGH, including considerations of privacy for data sharing.

