## Supplementary File for "Developing intrinsic capacity measures for research and practice in an Australian context"

### Appendix 1. Measurement methods for indicators of each intrinsic capacity domain

**Domain 1 - Cognition –** based on available literature and CHAMP data, the following indicators were considered:

1- Mini-Mental State Examination (MMSE) [1-3] (continuous, range: 0 – 30): MMSE comprises eleven questions and can be administered in 5-10 minutes. It primarily assesses mental functions, but not mood, abnormal mental experiences, and the form of thinking [4]. It contains 11 questions: 2 questions on orientation, 1 question on registration, 1 question on attention and calculation, 1 question on recall, 6 questions on language. A higher score represents better cognitive capacity.

2- Executive function measured based on TMT-B and Weigl Color Form Sorting [5-7]:

- The Trail Making Test (TMT) (binary: able to complete in 5 minutes, yes/no): it is used as an indicator of visual-motor skills and speed, and executive function [8]. It consists of two parts, TMT-A and TMT-B. In TMT-A, the respondent is asked to connect randomly arranged circled numbers; in TMT-B, the respondent is required to connect and alternate a numbers and letters sequence. The TMT-B has shown strength as a cognitive test and was included in CHAMP, and it was implemented in various studies as a measurement of cognition [1, 3, 6, 9]. The primary outcome of this task for our study was to determine if the participant could connect all 25 circles in five minutes [10].
- The Weigl Color Form Sorting Test (categorical: unable to do the test; unable to do the first sort; sorted one category spontaneously; sorts two categories spontaneously): it primarily examines executive functioning [11], participants were asked to sort the colour form pieces into separate groups.

4- Memory [3, 5, 12, 13] (continuous, range: 0 – 25 for both tasks) [14]: Immediate logical memory and delayed logical memory was evaluated using the Wechsler Memory Scale [15] as part of the cognitive assessment in CHAMP. For the immediate logical memory, participants are asked to immediately retell the story read by the interviewer, and for the delayed logical memory, participants are asked to retell the story read by the interviewer a while ago.

2- Verbal fluency [1, 2, 16] (continuous, range: 0 – 14): verbal fluency was assessed as a component of Addenbrooke's Cognitive Examination (ACE) [17] that measures phonemic fluency and semantic fluency by asking the participants to generate words from a specified letter of the alphabet and generate words of animals.

**Domain 2 - Vitality –** we adopted a set of indicators based on the literature review:

1 - Forced expiratory volume in 1 second (FEV1) (continuous: volume of air in litres): measured using a spirometer, the best result from the three tests was kept.

2 - Body mass index (BMI) (continuous: in kg/m^2^) [12, 18]. BMI was calculated using height and weight measured during clinical assessment.

3 - Hand grip strength (continuous: in kilograms): was measured using hand-held dynamometers.

4 - Weight loss (binary, yes/no): was defined as losing at least 15% lower weight than the self-reported heaviest weight, or weight at age 25 (retrospective recall) if the heaviest weight was unavailable [19].

5 - The waist-to-hip ratio (WHR) (continuous): it was included in addition to the commonly assessed mortality-related factor BMI, which has been shown to be positively associated with mortality and may offer additional insights [20].

**Domain 3: Locomotor function** – Studies commonly employed performance-based assessments for locomotor function through the following indicators:

1- Dynamic [21, 22] and static balance [12, 16, 22]:

- Dynamic balance (categorical: did test and had 2 or fewer deviations at least once; did at least one trial but had 3 or more deviations on all attempts; unable to do any trials): it was assessed by a 20-centimetre narrow walk.
- Static balance: the sway was measured using a sway meter as part of the static balance assessment (continuous for both sway tests). Participants were asked to stand still on the floor, followed by standing on a piece of foam rubber mat. The measurement outcomes were the total area in cm^2^ calculated by multiplying the vertical distance by the horizontal distance. Single chair stand (categorical: stands without using arms; rises using arms; unable to stand; did not attempt/refused) and five consecutive chair stand tasks (continuous: time of completion in seconds) were also conducted for measuring static balance.

2 - Walking and gait parameters [16, 21, 23]: Walking pace (continuous: pace in meters per second) was measured by a 6-meter walking test, in which participants were asked to walk at their normal pace, and walking aids were allowed.

3 - Muscle strength (continuous, in kilograms) [22]: was measured by a spring gauge (40kg or 100kg, depending on participants’ abilities).

4 - Hand grip strength (continuous, in kilograms): was measured using hand-held dynamometers. (Note: hand grip strength is considered for both locomotor function and Vitality).

**Domain 4 – Sensory –** Vision and hearing are commonly used measures of sensory function in studies [24-26], either through performance-based measures or self-reported assessments:

1 - Vision (continuous): Visual acuity was examined and recorded during clinical assessment, and visual impairment is defined as visual acuity of 20/40 or worse [27].

2 - Hearing loss (binary, yes/no): Participants responded to a question regarding whether they had experienced hearing loss in the CHAMP self-completed questionnaire.

**Domain 5 - Psychological well-being** – Indicators of psychological capacity are generally self-reported; we considered the following indicators based on existing studies and data availability in CHAMP data.

1 - Depression (binary, yes/no): was assessed using the 15-item Geriatric Depression Scale (GDS-15) [18, 28-30], it is a common tool for screening later-life depression and can reflect a stable core of individuals’ depressivity. The scale includes 15 yes-or-no questions regarding how individuals felt over the past week, participants who answered five or more “yes” were classified as “having depression” [31].

2 - Social satisfaction and social participation (continuous scores): were evaluated through the 11-item Duke Social Support Index (DSSI) [32].

3 - Anxiety (binary, yes/no) was assessed by the Goldberg Anxiety Scale (GAS) [33], men with a GAS score ≥ 5 were classified as experiencing clinical anxiety symptoms.

4 - Sleep problem and health worry were assessed through single binary questions from the Goldberg Anxiety Scale (GAS) [33].

### Appendix 2. Measurement and categories of socioeconomic indicators for describing baseline characteristics

Socioeconomic position was assessed using the highest education level, occupation category, source of income, and home ownership (owning home outright versus other arrangements, such as leasing or purchasing in a retirement village, paying rent to a private landlord, and paying rent to the government for public housing). The highest education level was classified as high (university degree), intermediate (trade, apprenticeship, certificate, or diploma), and low (no post-school qualification), broadly aligned with the International Standard Classification for Education [34, 35]. Occupation category was classified based on the longest occupation performed during working life and the Australian and New Zealand Standard Classification of Occupations, and was categorised into 3 groups: high (higher professionals and managers, lower professionals and managers, and higher clerical service), intermediate (small employers and self-employed, farmers, lower supervisors, and technicians), and low (lower clerical, service, sales workers, and skilled and unskilled workers) [36]. Source of income comprises 3 categories: high (sources of income do not include any government pension, intermediate (reliant on government pension plus other sources of income), and low (reliant solely on government pension).

### Appendix 3. Missing data

The percentages of missing data of indicators and relevant known reasons for missing responses included in the analysis are shown in the Supplementary Table 1. We used Little’s test to assess if the data were missing completely at random (MCAR). The test result indicates that the missingness was missing completely at random (p > 0.99).

Ignoring missing data risks biased estimates and loss of precision [37]. We therefore used multiple imputation, which is valid under missing at random and can incorporate information from incomplete cases and auxiliary variables to reduce bias and improve precision, in preference to complete-case analysis. The auxiliary variables included in multiple imputation are the number of children, age at baseline, country of birth, marital status, living alone, highest education level, occupation category, income source category, and house ownership. To further improve imputation, binary indicators capturing participants’ physical limitations, applied when specific assessments could not be completed, were generated and incorporated as auxiliary variables.

### Appendix 4. Rescaling indicators and scoring intrinsic capacity

To ensure consistent interpretation of the direction of estimated factor loadings, we recoded some numerical variables using min-max reverse rescaling so that higher values consistently indicated better capacity. For example, measures such as the area of postural sway, where larger values represent poorer stability, were reverse-scored. The same approach was applied to some binary variables by recoding in the same direction, and the reference category for each categorical indicator was specified as the level corresponding to the lowest performance or status.

Intrinsic capacity scores were computed as a weighted linear combination of observed indicators, using the factor score coefficient matrix and the standardised indicators. For each participant, the intrinsic capacity scores were computed as:

From the full model: $\hat{Intrnsic capacity}$ = *0.0837*MMSE score + 0.0739 (if completed TMT-B in 5 minutes) + 0.0396*logical memory + 0.0337*logical memory recall + 0.0916 *Verbal fluency + 0.1692*FEV1 - 0.0005*BMI + 0.0910*Handgrip strength + 0 .0583*reversed time of repeated chair stand + 0.0824* leg strength + 0.1664*walking pace + 0.0200*reversed floor sway area + 0.0308*Reversed foam sway area + 0.0398 (if completed single chair stand without arm) + 0.0087 (if completed single chair stand with arm) + 0.1456 (if had any success trial in narrow walk test) + 0.0830 (if unstable in narrow walk test) + 0.2199*visual acuity + 0.0063 (If no hearing loss) + 0.1391 (if no depression) + 0.0694*social interaction + 0.0664*social satisfaction + 0.0322 (if no anxiety)*

From the minimal-set model: $\hat{Intrnsic capacity}$ = *0.1240*MMSE score + 0.1238 (if completed TMT-B in 5 minutes) + 0.1181*FEV1 + 0.2312*Handgrip strength + 0.0852* reversed time of repeated chair stands + 0.3283*walking pace + 0.1882*visual acuity + 0.0072 (If no hearing loss) + 0.1255 (if no depression) + 0.0849*social interaction*

From the clinically informed model: $\hat{Intrnsic capacity}$ = *0.1269*MMSE score + 0.1025*logical memory + 0.1133*FEV1 + 0.2524*Handgrip strength + 0 .1025*reversed time of repeated chair stand + 0.3472*walking pace + 0.1780*visual acuity + 0.0099 (If no hearing loss) + 0.1396 (if no depression) + 0.0027 (if no anxiety)*

Intrinsic capacity scores derived from the CFA models were then standardised and rescaled to a 0–100 metric to facilitate interpretability and comparability across models.

### Appendix 5. Consultation with a geriatrician.

A list of all potential indicators relevant to the five domains of intrinsic capacity and available in the CHAMP data was reviewed by a geriatrician (VN), Professor of Geriatric Medicine at the University of Sydney, Consultant Geriatrician at Concord Hospital, and co-director of the Centre for Education and Research on Ageing (CERA). His clinical work involves patients in acute geriatric medicine and rehabilitation wards, as well as a monthly two-day clinic in regional New South Wales. As a past President of the Australian and New Zealand Society for Geriatric Medicine, he has a broad overview of clinical practice across the field. He was therefore well placed to assess the clinical utility of each indicator, drawing on both his direct patient care experience and his familiarity with practice patterns across the geriatric medicine community in Australia and New Zealand. For each indicator, he was asked to provide a binary response (yes/no) regarding its routine use and clinical utility in geriatric practice, along with written comments to contextualise his assessment. Indicators endorsed as commonly used and clinically meaningful for assessing intrinsic capacity were included in the clinically informed CFA model.

### Appendix 6. Item response theory models and results

To simplify interpretation, we coded continuous variables (MMSE score, logical memory, logical memory recall and verbal fluency in cognition Domain; handgrip, FEV1 and BMI in vitality domain, walking pace, repeated chair stand, reversed floor sway area, reversed foam sway area, handgrip and leg strength in locomotor function domain, visual acuity in sensory domain, social interaction and social satisfaction in psychological well-being domain) into quartiles using the ntile function in R.

- Model specification: “graded” (graded response model) for ordinal variables included the categorical indicators and categorised continuous indicators mentioned above

And

“2pl” (2 parameter logistic model) for binary indicators.

- Estimation method: standard Expectation-Maximisation algorithm with fixed quadrature.

Item information curves are shown in the Figure below.

### Appendix 6, Figure. Item information curves of the item response theory model.

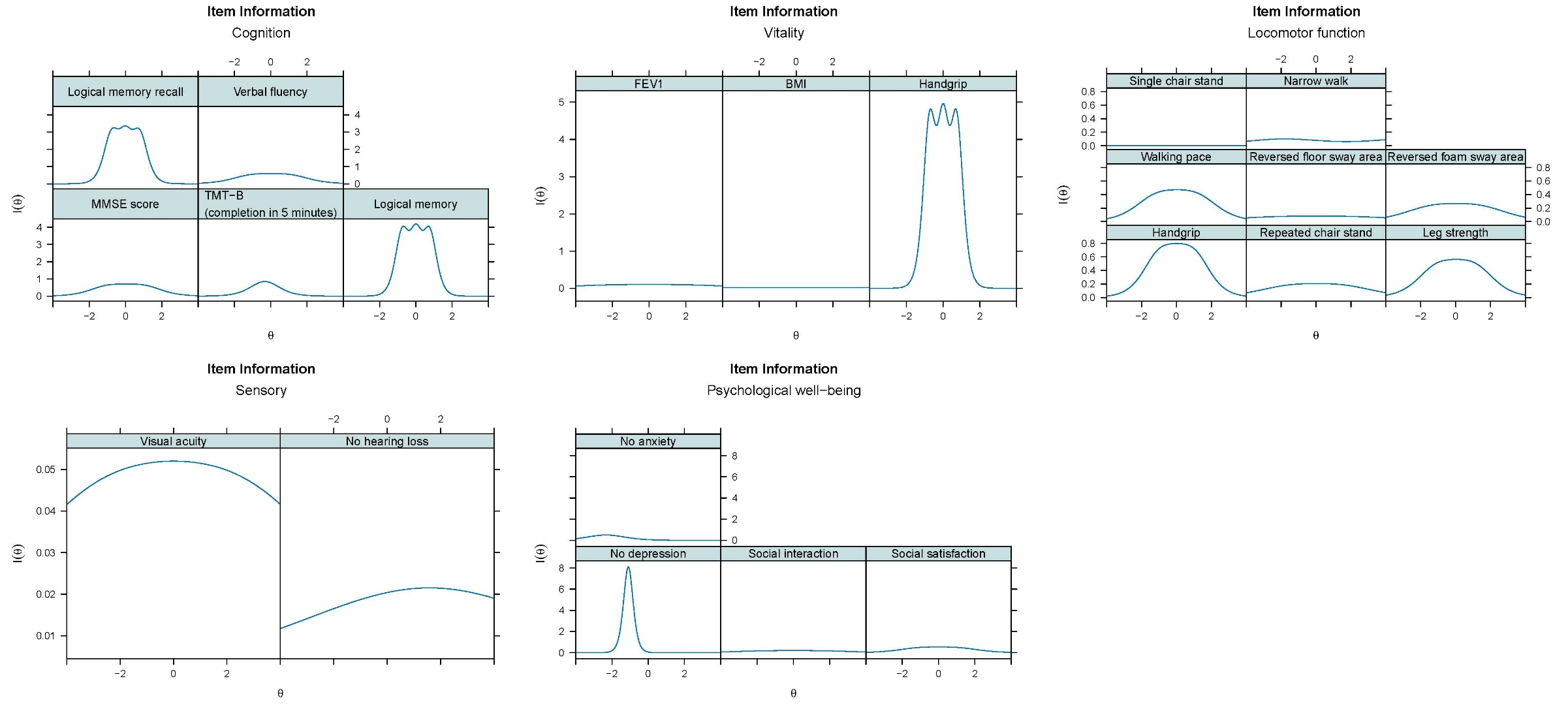

Note: Continuous indicators were categorised into quartiles.

The item information curves show the ability for each item to precisely quantify an individual’s intrinsic capacity domain, across the range of each domain of intrinsic capacity. Results show that most domains contained at least one item with strong information near the centre of the intrinsic capacity domain distribution, where the majority of the population is expected to fall (<5% of the population expected to have a standardised intrinsic capacity of <-2 or >2). Locomotor function items showed comparatively lower information overall relative to other domains, while Sensory items (e.g., visual acuity) showed broader but comparatively low-magnitude information across the range. Psychological well-being items, by contrast, showed higher information below the mean but markedly less above it. Taken together, these results suggest that for most domains, no single item provides precise measurement across the full range of intrinsic capacity, and combining multiple items is therefore necessary to adequately capture the whole distribution.

### Table S1. STROBE Statement – Checklist of items that should be included in reports of cohort studies

|  | Item No | Recommendation | Page No |
| --- | --- | --- | --- |
| **Title and abstract** | 1 | (*a*) Indicate the study’s design with a commonly used term in the title or the abstract | 1-3 |
|  |  | (*b*) Provide in the abstract an informative and balanced summary of what was done and what was found | 3 |
| Introduction | | | |
| Background/rationale | 2 | Explain the scientific background and rationale for the investigation being reported | 3-4 |
| Objectives | 3 | State specific objectives, including any prespecified hypotheses | 3-4 |
| Methods | | | |
| Study design | 4 | Present key elements of study design early in the paper | 4-5 |
| Setting | 5 | Describe the setting, locations, and relevant dates, including periods of recruitment, exposure, follow-up, and data collection | 5-6 |
| Participants | 6 | (*a*) Give the eligibility criteria, and the sources and methods of selection of participants. Describe methods of follow-up | 5 |
|  |  | (*b*) For matched studies, give matching criteria and number of exposed and unexposed | Not applicable |
| Variables | 7 | Clearly define all outcomes, exposures, predictors, potential confounders, and effect modifiers. Give diagnostic criteria, if applicable | 5-7 |
| Data sources/ measurement | 8 | For each variable of interest, give sources of data and details of methods of assessment (measurement). Describe comparability of assessment methods if there is more than one group | 5-7;  Supplementary Material pages 2-4 and 12-14 |
| Bias | 9 | Describe any efforts to address potential sources of bias | 12-14 |
| Study size | 10 | Explain how the study size was arrived at | 5 |
| Quantitative variables | 11 | Explain how quantitative variables were handled in the analyses. If applicable, describe which groupings were chosen and why | 6-7 |
| Statistical methods | 12 | (*a*) Describe all statistical methods, including those used to control for confounding | 6-8 |
|  |  | (*b*) Describe any methods used to examine subgroups and interactions | Not applicable |
|  |  | (*c*) Explain how missing data were addressed | 7;  Supplementary Materials page 7 (Appendix 3) |
|  |  | (*d*) If applicable, explain how loss to follow-up was addressed | 5; Figure S1 |
|  |  | (*e*) Describe any sensitivity analyses | 7-9 |
| Results | | |  |
| Participants | 13 | (a) Report numbers of individuals at each stage of study—eg numbers potentially eligible, examined for eligibility, confirmed eligible, included in the study, completing follow-up, and analysed | 5, 7 |
|  |  | (b) Give reasons for non-participation at each stage | Not applicable |
|  |  | (c) Consider use of a flow diagram | Supplementary Material page 18. (Supplementary Figure S1) |
| Descriptive data | 14 | (a) Give characteristics of study participants (eg demographic, clinical, social) and information on exposures and potential confounders | 5, 10 and Table 1 (page 19-20) |
|  |  | (b) Indicate number of participants with missing data for each variable of interest | Supplementary Material page 12-13 |
|  |  | (c) Summarise follow-up time (eg, average and total amount) | 9 |
| Outcome data | 15 | Report numbers of outcome events or summary measures over time | 9-11 |
| Main results | 16 | (*a*) Give unadjusted estimates and, if applicable, confounder-adjusted estimates and their precision (eg, 95% confidence interval). Make clear which confounders were adjusted for and why they were included | 9-11 |
|  |  | (*b*) Report category boundaries when continuous variables were categorized | Not applicable |
|  |  | (*c*) If relevant, consider translating estimates of relative risk into absolute risk for a meaningful time period | 11 |
| Other analyses | 17 | Report other analyses done—eg analyses of subgroups and interactions, and sensitivity analyses | None |
| **Discussion** |  |  |  |
| Key results | 18 | Summarise key results with reference to study objectives | 10-11 |
| Limitations | 19 | Discuss limitations of the study, taking into account sources of potential bias or imprecision. Discuss both direction and magnitude of any potential bias | 11-14 |
| Interpretation | 20 | Give a cautious overall interpretation of results considering objectives, limitations, multiplicity of analyses, results from similar studies, and other relevant evidence | 10-14 |
| Generalisability | 21 | Discuss the generalisability (external validity) of the study results | 13-14 |
| **Other information** |  |  |  |
| Funding | 22 | Give the source of funding and the role of the funders for the present study and, if applicable, for the original study on which the present article is based | 1 |

### Table S2. Measurement methods, instruments, and missing data for intrinsic capacity domain indicators

| Intrinsic Capacity Domains | Domain Indicators | Instrument | Missing data at baseline (n, %) | Relevant inabilities | Task-irrelevant reasons of being unable |
| --- | --- | --- | --- | --- | --- |
| Cognition | MMSE | MMSE [4] | 186 (10.9%) | Dementia; dyslexia; Alzheimer disease; CVA (cerebrovascular accident); stroke; head injury; neuropathy; confused; conscious  (n = 16; 0.9%) | Language barriers;  Other reasons:  visual impairment; hearing problems; tremors; Parkinson’s disease; breathless; dyspnoea; other physical disability; surgeries irrelevant to cognitive function) |
|  | Executive function | TMT-B [8, 38] | 8 (0.5%) | -- |  |
|  |  | Weigl color form sorting test [11] | 143 (8.4%) | Reasons same as above  (n = 3; 0.2%) |  |
|  | Memory | Logical memory (immediate) | 229 (13.4%) | Reasons same as above  (n = 41; 2.4%) |  |
|  | Delayed memory | Logical memory (recall) | 229 (13.4%) | Reasons same as above  (n = 41; 2.4%) |  |
|  | Verbal fluency | 2 questions from the ACE [17] | 282 (16.5%) | Reasons same as above  (n = 43; 2.5%) |  |
| Vitality | Force expiratory volume (FEV1) | FEV1 | 413 (24.2%) | The test was not conducted if any of the following conditions existed:  Surgery on chest or abdomen in the past three months; heart attack within the past three months; detached retina or eye surgery in the past three months; hospitalised due to heart problems in the past three months; resting pulse > 120 best per minute  (n = 132; 7.7%) | Unable to perform adequate test; did not understand instructions |
|  | Body mass index | -- | 28 (1.6%) | N/A | -- |
|  | Handgrip strength | -- | 101 (5.9%) | The test was not conducted if any of the following conditions existed:  Pain or arthritis in both hands; surgery on both hands or wrists in the past three months  (n = 92; 5.4%) | -- |
|  | Weight loss | -- | 55 (3.2%) | N/A | -- |
|  | Waist-to-hip ratio | -- | 42 (2.5%) | N/A | N/A |
| Locomotor function | Dynamic balance | Single chair stand | 64 (3.8%) | Physical inability | -- |
|  |  | Repeated chair stand | 151 (8.9%) | Physical inability; could not stand without using arm in the single-sit-to-stand task  (n = 113; 6.6%) | -- |
|  |  | 20 cm narrow walk | 23 (1.3%) | N/A | -- |
|  | Static balance | Floor sway area | 88 (5.2%) | Physical inability  (n = 60; 3.5%) | -- |
|  |  | Foam sway area | 205 (12.0%) | Physical inability  (n = 164; 9.6%) | -- |
|  | Walking and gait parameters | 6-meter walk pace | 77 (4.5%) | Physical inability  (n = 41; 2.4%) | -- |
| Sensory | Visual acuity | LogMAR chart | 109 (6.4%) | Blindness; macular degeneration  (n = 16; 0.9%) | Language barriers; cognitive limitations |
|  | Hearing loss (yes/no) | -- | 23 (1.3%) | -- | -- |
| Psychological wellbeing | Depression | 15-item Geriatric Depression Scale (GDS-15) [39] | 24 (1.4%) | N/A | N/A |
|  | Life satisfaction | Duke Social Support Index [32] – Satisfaction | 23 (1.3%) | N/A | N/A |
|  | Social participation | Duke Social Support Index [32]– Interaction | 40 (2.3%) | N/A | N/A |
|  | Social network | Duke Social Support Index [32] – Network | 33 (1.9%) | N/A | N/A |
|  | Anxiety | Goldberg Anxiety Scale [33] | 34 (2%) | N/A | N/A |
|  | Sleep problem | Goldberg Anxiety Scale [33] | 37 (2.2%) | N/A | N/A |
|  | Health worries | Goldberg Anxiety Scale [33] | 33 (1.9%) | N/A | N/A |

Abbreviations: MMSE, Mini-Mental State Examination; TMT, Trail Making Test; FEV1, forced expiratory volume in 1 second; BMI, body mass index.

### Table S3. Activities of daily living (ADL) and instrumental activities of daily living (IADL) scales

| ADL items [40] | |
| --- | --- |
| “Do you need help from another person or special equipment or device to do any of the following things?” | |
| Walking across a small room? | No, does not need help |
|  | Yes, needs help |
|  | Unable to do this |
| Bathing, either a sponge bath, tub bath, or shower? | No, does not need help |
|  | Yes, needs help |
|  | Unable to do this |
| Personal grooming, like brushing hair, brushing teeth, or washing face? | No, does not need help |
|  | Yes, needs help |
|  | Unable to do this |
| Dressing, like putting on a shirt, buttoning and zipping, or putting on shoes? | No, does not need help |
|  | Yes, needs help |
|  | Unable to do this |
| Eating like holding a fork, cutting food, or drinking from a glass? | No, does not need help |
|  | Yes, needs help |
|  | Unable to do this |
| Getting from a bed to a chair? | No, does not need help |
|  | Yes, needs help |
|  | Unable to do this |
| Using the toilet? | No, does not need help |
|  | Yes, needs help |
|  | Unable to do this |
| IADL items [41] | |
| “Can you can do these activities without any help at all, or if you need some help to do them, or if you can’t do them at all?” | |
| Can you use the telephone? | Without help |
|  | With some help |
|  | Completely unable |
| Can you get to places out of walking distance? | Without help |
|  | With some help |
|  | Completely unable |
| Can you go shopping for groceries or clothes (if you have transportation)? | Without help |
|  | With some help |
|  | Completely unable |
| Can you prepare you own meals? | Without help |
|  | With some help |
|  | Completely unable |
| Can you do your housework? | Without help |
|  | With some help |
|  | Completely unable |
| Can you take your own medications? | Without help |
|  | With some help |
|  | Completely unable |
| Can you handle your own money? | Without help |
|  | With some help |
|  | Completely unable |
| Are you able to do heavy work around the house, like washing windows, walls, or floors without help? | Yes |
|  | No |
| Are you able to walk up and down stairs to the first floor without help? | Yes |
|  | No |
| Are you able to walk half a mile (approximately one kilometre) without help? | Yes |
|  | No |

### Table S4. Confirmatory factor analysis models and related root mean square error of approximation (RMSEA) to identify a simplified model that best reflects intrinsic capacity

| **Model iterations** | | **1** | **2** | **3** | **4** | **5** | **6** | **7** | **8** | **9** | **10** | **11** | **12** | **13** | **14** | **15** | **16** | **17** | **18** | **19** | **20** | **21** |
| --- | --- | --- | --- | --- | --- | --- | --- | --- | --- | --- | --- | --- | --- | --- | --- | --- | --- | --- | --- | --- | --- | --- |
| Cognition | MMSE | ✓ | ✓ | ✓ | ✓ | ✓ | ✓ | ✓ | ✓ | ✓ | ✓ | ✓ | ✓ | ✓ | ✓ | ✓ | ✓ | ✓ | ✓ | ✓ | ✓ | ✓ |
|  | TMT-B | ✓ | ✓ | ✓ | ✓ | ✓ | ✓ | ✓ | ✓ | ✓ | ✓ | ✓ | ✓ | ✓ | ✓ | ✓ | ✓ | ✓ | ✓ | ✓ | ✓ | ✓ |
|  | Logical memory | ✓ | ✓ | ✓ | ✓ | ✓ | ✓ | ✓ | ✓ | ✓ | ✓ |  |  | ✓ | ✓ | ✓ | ✓ | ✓ | ✓ | ✓ | ✓ | ✓ |
|  | Logical memory recall | ✓ | ✓ | ✓ | ✓ | ✓ | ✓ | ✓ | ✓ | ✓ |  |  |  | ✓ | ✓ | ✓ | ✓ | ✓ | ✓ | ✓ | ✓ |  |
|  | Verbal fluency | ✓ | ✓ | ✓ | ✓ | ✓ | ✓ | ✓ | ✓ | ✓ | ✓ | ✓ |  | ✓ | ✓ | ✓ | ✓ | ✓ | ✓ | ✓ |  |  |
| Vitality | FEV1 | ✓ | ✓ | ✓ | ✓ | ✓ | ✓ | ✓ | ✓ | ✓ | ✓ | ✓ | ✓ | ✓ | ✓ | ✓ | ✓ | ✓ | ✓ | ✓ | ✓ | ✓ |
|  | BMI | ✓ |  |  |  |  |  |  |  |  |  |  |  | ✓ | ✓ | ✓ | ✓ | ✓ | ✓ | ✓ | ✓ | ✓ |
|  | Best grip | ✓ | ✓ | ✓ | ✓ | ✓ | ✓ | ✓ | ✓ | ✓ | ✓ | ✓ | ✓ | ✓ | ✓ | ✓ | ✓ | ✓ | ✓ | ✓ | ✓ | ✓ |
| Locomotor function  Sensory | Repeated chair stand | ✓ | ✓ | ✓ | ✓ | ✓ | ✓ | ✓ | ✓ | ✓ | ✓ | ✓ | ✓ | ✓ | ✓ | ✓ | ✓ | ✓ | ✓ | ✓ | ✓ | ✓ |
|  | Knee strength | ✓ | ✓ | ✓ | ✓ | ✓ | ✓ |  |  |  |  |  |  | ✓ | ✓ | ✓ | ✓ |  |  |  |  |  |
|  | Walk pace (6m) | ✓ | ✓ | ✓ | ✓ | ✓ | ✓ | ✓ | ✓ | ✓ | ✓ | ✓ | ✓ | ✓ | ✓ | ✓ |  |  |  |  |  |  |
|  | Floor sway | ✓ | ✓ | ✓ |  |  |  |  |  |  |  |  |  | ✓ | ✓ | ✓ | ✓ | ✓ | ✓ | ✓ | ✓ | ✓ |
|  | Foam sway | ✓ | ✓ | ✓ | ✓ | ✓ |  |  |  |  |  |  |  | ✓ | ✓ | ✓ | ✓ | ✓ | ✓ |  |  |  |
|  | Single chair stand | ✓ | ✓ |  |  |  |  |  |  |  |  |  |  | ✓ | ✓ | ✓ | ✓ | ✓ |  |  |  |  |
|  | Narrow walk | ✓ | ✓ | ✓ | ✓ | ✓ | ✓ | ✓ |  |  |  |  |  | ✓ | ✓ | ✓ | ✓ | ✓ | ✓ | ✓ | ✓ | ✓ |
|  | Best grip | ✓ | ✓ | ✓ |  |  |  |  |  |  |  |  |  | ✓ | ✓ |  |  |  |  |  |  |  |
| Sensory | Acuity | ✓ | ✓ | ✓ | ✓ | ✓ | ✓ | ✓ | ✓ | ✓ | ✓ | ✓ | ✓ | ✓ | ✓ | ✓ | ✓ | ✓ | ✓ | ✓ | ✓ | ✓ |
|  | Hearing loss | ✓ | ✓ | ✓ | ✓ | ✓ | ✓ | ✓ | ✓ | ✓ | ✓ | ✓ | ✓ | ✓ | ✓ | ✓ | ✓ | ✓ | ✓ | ✓ | ✓ | ✓ |
| Psychological wellbeing | Depression | ✓ | ✓ | ✓ | ✓ | ✓ | ✓ | ✓ | ✓ | ✓ | ✓ | ✓ | ✓ |  |  |  |  |  |  |  |  |  |
|  | Social interaction | ✓ | ✓ | ✓ | ✓ | ✓ | ✓ | ✓ | ✓ | ✓ | ✓ | ✓ | ✓ | ✓ |  |  |  |  |  |  |  |  |
|  | Social satisfaction | ✓ | ✓ | ✓ | ✓ | ✓ | ✓ | ✓ | ✓ |  |  |  |  | ✓ | ✓ | ✓ | ✓ | ✓ | ✓ | ✓ | ✓ | ✓ |
|  | Anxiety | ✓ | ✓ | ✓ | ✓ |  |  |  |  |  |  |  |  | ✓ | ✓ | ✓ | ✓ | ✓ | ✓ | ✓ | ✓ | ✓ |
| Robust RMSEA | | 0.026 | 0.018 | 0.022 | 0.026 | 0.027 | 0.028 | 0.023 | 0.016 | 0.016 | 0.018 | 0.021 | 0.024 | 0.025 | 0.026 | 0.031 | 0.031 | 0.025 | 0.028 | 0.030 | 0.030 | 0.032 |
| Robust CFI | | 0.974 | 0.994 | 0.990 | 0.982 | 0.976 | 0.977 | 0.986 | 1.000 | 0.995 | 0.998 | 0.995 | 0.990 | 0.967 | 0.977 | 0.962 | 0.951 | 0.969 | 0.978 | 0.973 | 0.975 | 0.972 |
| Robust TLI | | 0.969 | 0.993 | 0.988 | 0.978 | 0.971 | 0.972 | 0.983 | 1.000 | 0.993 | 0.997 | 0.993 | 0.986 | 0.961 | 0.972 | 0.955 | 0.941 | 0.963 | 0.972 | 0.966 | 0.969 | 0.964 |
| SRMR | | 0.037 | 0.033 | 0.032 | 0.035 | 0.033 | 0.035 | 0.029 | 0.025 | 0.023 | 0.024 | 0.025 | 0.025 | 0.036 | 0.036 | 0.039 | 0.039 | 0.032 | 0.032 | 0.030 | 0.033 | 0.034 |

| **Model iterations** | | **22** | **23** | **24** | **25** | **26** | **27** | **28** | **29** | **30** | **31** | **32** | **33** | **34** |
| --- | --- | --- | --- | --- | --- | --- | --- | --- | --- | --- | --- | --- | --- | --- |
| Cognition | MMSE | ✓ | ✓ | ✓ | ✓ | ✓ | ✓ | ✓ |  | ✓ | ✓ | ✓ | ✓ |  |
|  | TMT-B | ✓ |  |  | ✓ |  | ✓ | ✓ | ✓ | ✓ | ✓ | ✓ | ✓ | ✓ |
|  | Logical memory |  | ✓ | ✓ |  | ✓ |  |  |  |  |  |  |  |  |
|  | Logical memory recall |  |  |  |  |  |  |  |  |  |  |  |  |  |
|  | Verbal fluency |  |  |  |  |  |  |  | ✓ |  |  |  |  | ✓ |
| Vitality | FEV1 | ✓ | ✓ |  | ✓ | ✓ | ✓ | ✓ | ✓ | ✓ | ✓ | ✓ | ✓ | ✓ |
|  | BMI |  |  |  |  |  |  |  |  |  |  |  |  |  |
|  | Best grip | ✓ | ✓ | ✓ | ✓ | ✓ | ✓ | ✓ | ✓ | ✓ | ✓ | ✓ | ✓ | ✓ |
| Locomotor function  Sensory | Repeated chair stand | ✓ | ✓ | ✓ | ✓ | ✓ |  |  |  |  | ✓ | ✓ |  |  |
|  | Knee strength |  |  |  |  |  | ✓ |  |  |  |  |  |  |  |
|  | Walk pace (6m) |  | ✓ | ✓ | ✓ | ✓ | ✓ | ✓ | ✓ | ✓ | ✓ | ✓ | ✓ | ✓ |
|  | Floor sway |  |  |  |  |  |  |  |  |  |  |  |  |  |
|  | Foam sway |  |  |  |  |  |  |  |  |  |  |  |  |  |
|  | Single chair stand |  |  |  |  |  |  |  |  |  |  |  |  |  |
|  | Narrow walk | ✓ |  |  |  |  |  | ✓ | ✓ | ✓ |  |  | ✓ | ✓ |
|  | Best grip |  |  |  |  |  |  |  |  |  |  |  |  |  |
| Sensory | Acuity | ✓ | ✓ | ✓ | ✓ | ✓ | ✓ | ✓ | ✓ | ✓ | ✓ | ✓ | ✓ | ✓ |
|  | Hearing loss | ✓ | ✓ | ✓ | ✓ | ✓ | ✓ | ✓ | ✓ | ✓ | ✓ | ✓ | ✓ | ✓ |
| Psychological wellbeing | GDS depression |  | ✓ | ✓ | ✓ | ✓ | ✓ | ✓ | ✓ | ✓ | ✓ | ✓ | ✓ | ✓ |
|  | Social interaction |  |  |  |  | ✓ | ✓ | ✓ |  |  |  |  |  |  |
|  | Social satisfaction | ✓ |  |  | ✓ |  |  |  | ✓ | ✓ | ✓ |  |  |  |
|  | Anxiety | ✓ | ✓ | ✓ |  |  |  |  |  |  |  | ✓ | ✓ | ✓ |
| Robust RMSEA | | 0.029 | 0.019 | 0.020 | 0.024 | 0.017 | 0.040 | 0.028 | 0.030 | 0.028 | 0.028 | 0.026 | 0.028 | 0.029 |
| Robust CFI | | 0.980 | 0.994 | 0.987 | 0.985 | 0.991 | 0.975 | 0.990 | 0.984 | 0.986 | 0.998 | 0.986 | 0.985 | 0.985 |
| Robust TLI | | 0.970 | 0.991 | 0.980 | 0.977 | 0.986 | 0.963 | 0.985 | 0.977 | 0.979 | 0.988 | 0.978 | 0.978 | 0.978 |
| SRMR | | 0.029 | 0.024 | 0.025 | 0.026 | 0.023 | 0.036 | 0.028 | 0.029 | 0.028 | 0.013 | 0.027 | 0.029 | 0.028 |

| Colour coding of the intrinsic capacity models |
| --- |
| Literature-based |
| Minimum-set |
| Clinically informed |

Abbreviations: MMSE, Mini-Mental State Examination; TMT, Trail Making Test; FEV1, forced expiratory volume in 1 second; BMI, body mass index; RMSEA: root mean square error of approximation; CFI: Comparative fit index; TLI: Tucker–Lewis index; SRMR: Standardised root mean square residual.

Model 1 is the comprehensive confirmatory factor analysis (CFA) model (Literature-based model).

Models 2 to 12 were obtained by excluding indicators with the lowest factor loading.

Model 12 is the final simplified CFA model (Minimum-set model).

Models 13 to 22 were obtained by excluding indicators with the highest standard error.

Models 23 and 24 are based on indicators suggested by a geriatrician for clinical applicability. Model 23 was adopted as the clinically informed model.

Models 25 to 28 were obtained by replacing indicators in the final two-indicator model with alternative indicators that had been excluded during model simplification due to lower factor loadings.

Models 29 to 34 were obtained by including combinations of the indicators with the top factor loadings in the domain-specific CFA models or in the initial comprehensive model, with consideration of distinctiveness and content coverage of the indicators.

### Figure S1. Flow diagram of included participants from the CHAMP study.

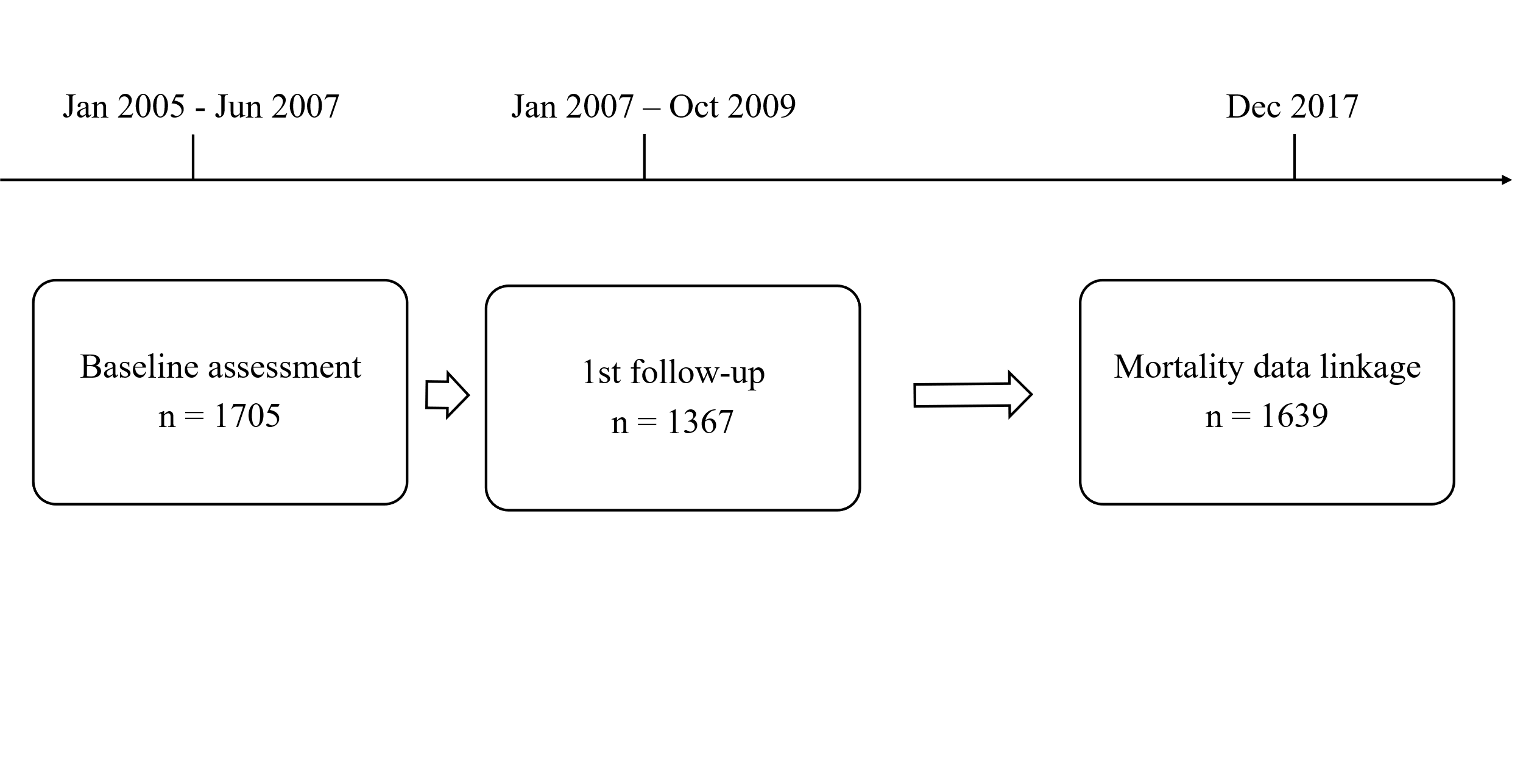

Of the original sample, 66 participants did not consent to mortality data linkage, hence n=1,639.

### Figure S2. Histograms of intrinsic capacity scores.

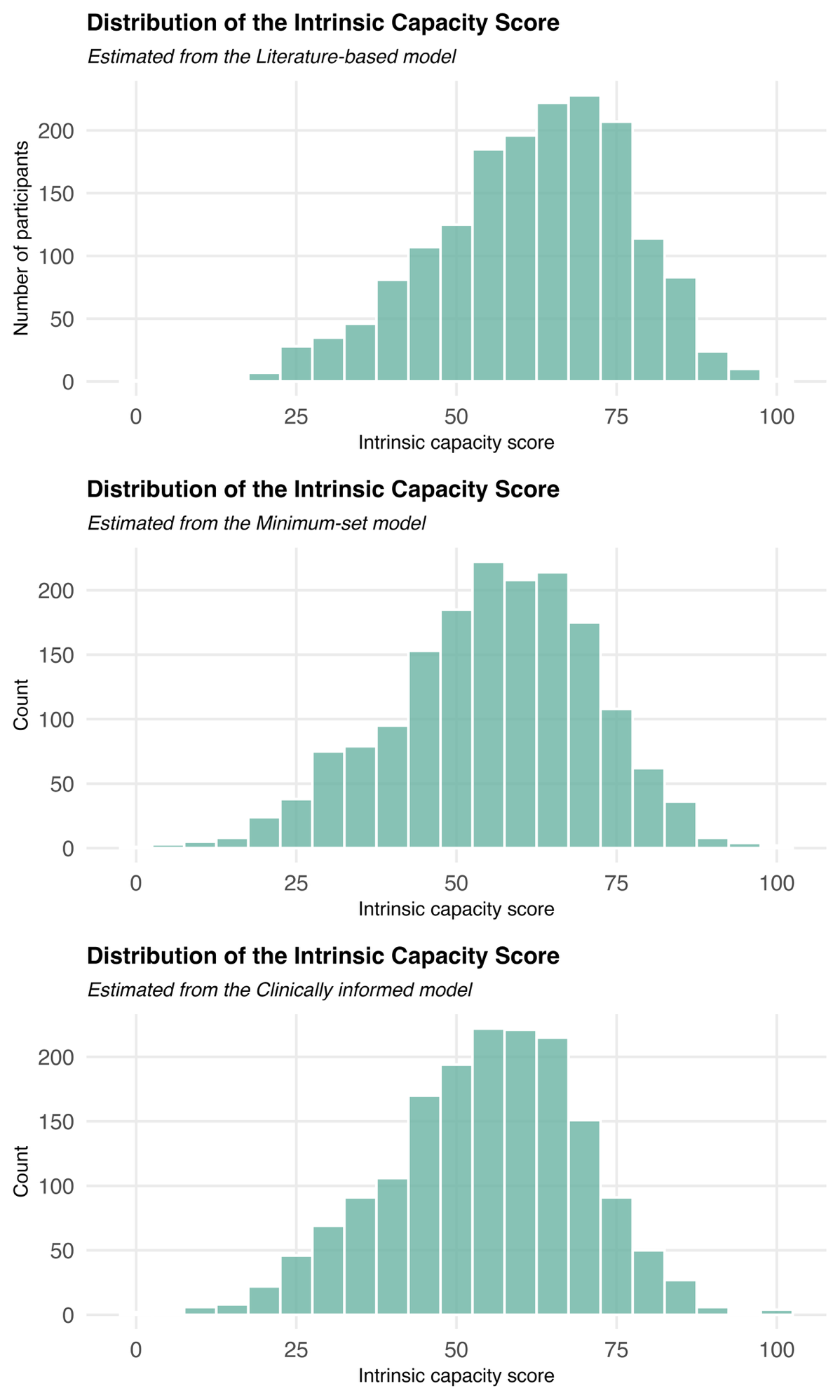
